# Serum Raman spectroscopy as a blood-based triage test for bladder cancer: a retrospective proof-of-concept study

**DOI:** 10.64898/2026.10.26.26366501

**Authors:** Ian Smith, Kirsty Milligan, Anton Flugge, Nuha Fakhrua Aldeen, Brian Shine, Cecilia Muldoon

## Abstract

**Background:** Bladder cancer remains one of the most common urological malignancies. Current diagnostic pathways rely heavily on cystoscopy, an invasive investigation with significant cost and resource implications. A rapid blood-based assay capable of improving patient stratification before cystoscopy could reduce unnecessary invasive investigations while maintaining diagnostic sensitivity.

**Methods:** Archived serum samples from patients with histologically confirmed bladder cancer and symptomatic control subjects were analysed using dried-serum Raman spectroscopy. Raman spectra were incorporated into supervised machine-learning classification models to generate predicted probabilities of bladder cancer. Diagnostic performance was evaluated using receiver operating characteristic (ROC) analysis.

**Results:** The study included 49 patients with bladder cancer and 97 symptomatic controls. Predicted probabilities of bladder cancer were substantially higher in cancer patients than in controls. The Raman classifier demonstrated excellent diagnostic discrimination (AUC approximately 0.90). Incorporation of age into the classification model produced little additional improvement (AUC approximately 0.91). The diagnostic test produced sensitivity and specificity values of 92% and 83%, respectively.

**Conclusions:** Machine learning-assisted analysis of dried-serum Raman spectra demonstrates promising diagnostic performance for bladder cancer. Prospective evaluation is warranted to determine its role as a minimally invasive triage test within contemporary bladder cancer diagnostic pathways.

## Introduction

Patients presenting with visible or non-visible haematuria represent a common and important diagnostic challenge in urological practice. Although most individuals do not have malignancy, haematuria may be the presenting feature of cancers arising anywhere within the urinary tract, including the kidney, renal pelvis, ureter and bladder.[1]Consequently, current clinical pathways aim to identify those patients requiring definitive investigation while avoiding unnecessary invasive procedures.

The subsequent investigation depends upon the suspected site of disease. Cross-sectional imaging plays the principal role in the diagnosis of renal malignancy, [2] whereas cystoscopy remains the reference investigation for bladder cancer. [1] Upper tract urothelial tumours are uncommon but may require dedicated imaging and ureteroscopy when clinically suspected.

Despite advances in imaging and endoscopy, there remains interest in minimally invasive laboratory tests capable of improving risk stratification at an earlier stage of the diagnostic pathway. Although several urinary biomarkers have been developed for bladder cancer, [3] [4] [5] [6] [7] [8] [9] [10] no blood-based laboratory test has become established in routine clinical practice.

Raman spectroscopy is a vibrational spectroscopic technique that provides a molecular fingerprint of biological samples without requiring targeted molecular assays. [11] Rather than measuring predefined biomarkers, Raman spectroscopy interrogates the composite biochemical composition of serum, potentially capturing disease-associated molecular changes distributed across multiple proteins, lipids and metabolites. [12] [13] [14]

Recent advances in instrumentation and machine-learning methods have renewed interest in Raman spectroscopy as a clinical diagnostic tool. [15] [16] [17] [12] Several studies have demonstrated its potential for distinguishing malignant from non-malignant disease, [18] [13] [19] [19] [20] [21] although relatively few have examined how serum Raman assays might be incorporated into established clinical pathways for bladder cancer.

The present proof-of-concept study evaluated a machine learning-assisted dried-serum Raman assay using archived serum samples from patients investigated for bladder cancer. [22] [23] We examined whether serum Raman spectroscopy could discriminate patients with bladder cancer from symptomatic controls and explored its potential role as a blood-based triage test within contemporary diagnostic pathways.

## Methods

### Study design

This retrospective proof-of-concept study evaluated the performance of a machine learning-assisted serum Raman spectroscopy assay for discriminating bladder cancer from non-malignant urological disease.

### Clinical Trial Number

Not applicable in this study.

### Patient cohort

Serum samples were obtained from the QUAlity biobanking iNvesTigating Urological Malignancies (QUANTUM) biobank, Nuffield Department of Surgical Sciences, University of Oxford. [22]

The study cohort comprised 152 participants, including 51 patients with histologically confirmed bladder cancer and 101 symptomatic control subjects without evidence of bladder cancer. The details of the cohort used in this study are given in Table 1.

**Table 1.** Cohort details.

|  | **Controls** | **Bladder Cancer** |
|---|---|---|
| **Average age (range)** | 65 (48-79) | 70 (44-87) |
| **Sex (m/f)** | (101/0) | (43/8) |
| **T Stage** | - | Ta (25) of which CIS (2)<br>T1 (15) of which CIS (1)<br>T2 (5)<br>Not stated (6) |
| **Grade** | - | G1 (3)<br>G2 (17)<br>G3 (14)<br>PUNLMP (4)<br>Not stated (13) |
| **Tumour type** | - | TCC (23)<br>Other (14)<br>Not stated (14) |
| **Growth pattern** | - | Papillary (27)<br>Solid (5)<br>Other (2)<br>Not Stated (17) |

### Serum preparation

Frozen serum samples were thawed immediately before analysis. Ten microlitres of serum were deposited onto aluminium foil mounted on a glass microscope slide and allowed to dry under ambient laboratory conditions before Raman spectral acquisition. The workflow is depicted in **Figure 1** Dried serum Raman analysis workflow

**Figure 1.**
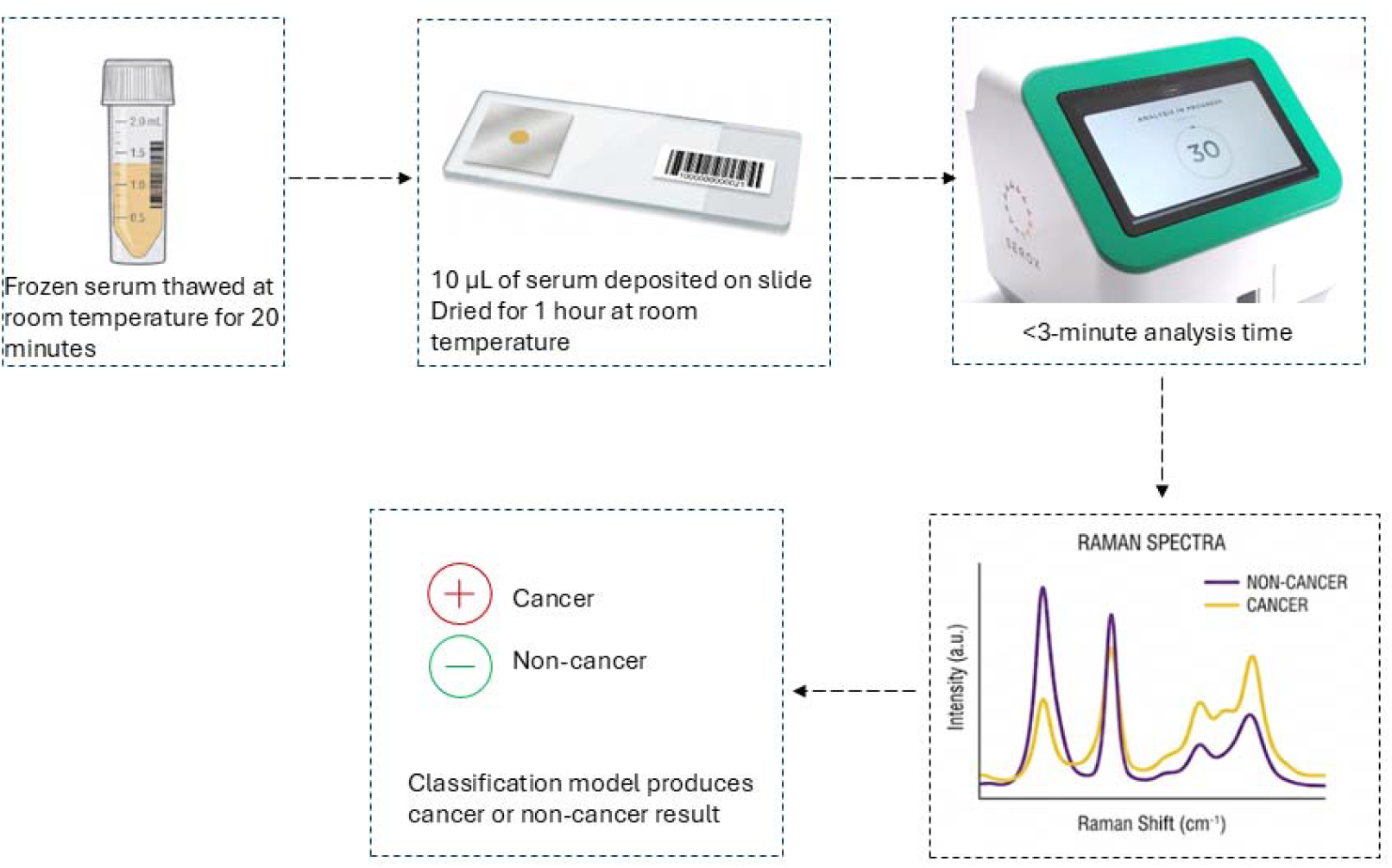
Dried serum Raman analysis workflow

### Raman spectroscopy

Spectra were acquired using a Wasatch Photonics WP-785 Raman spectrometer equipped with a 785 nm excitation laser. [24] Experimental parameters were defined as described in **Table 2**.

**Table 2.** Raman acquisition parameters.

| **Spectrometer model** | WP-785-R-SR-LMMFC-IC |
|---|---|
| **Excitation wavelength** | 785 nm |
| **Laser power** | 450 mW, multimode |
| **Acquisition time** | 500 ms |
| **Laser spot size (nomi-nal)** | 120 µm |
| **Aperture** | f/1.3 |
| **Slit size** | 25 µm |
| **Resolution** | 7 cm<sup>-1</sup> |
| **Spectral range** | 270-2700 cm<sup>-1</sup> |

### Spectral preprocessing and data exclusion

The signal to noise ratio of each spectrum was evaluated by computing the ratio of four defined peaks in the spectrum with noise in a silent region of the spectrum. As a result of this filtering step, spectra from four control samples and two bladder cancer samples (male) were excluded because of insufficient signal to noise ratio. The remaining 146 spectra were L2 normalised.

### Machine-learning analysis

Processed spectra were incorporated into a supervised machine-learning classification model to generate predicted probability of bladder cancer.

Two models were evaluated:

1. Raman spectra alone.
2. Raman spectra combined with patient age.

Both models were based on a gradient boosting tree model (XGBoost) [29] and evaluated using five-fold cross-validation. The output of the models are scores which represent the likelihood of each sample to belong to the bladder cancer cohort (probability score). Principal component analysis (PCA) was employed to reduce the dimensionality of the spectra. The model was tested 100 times using different random folds each time, to ensure robustness of output. All reported results are the average of 100 repetitions of the model, unless stated otherwise.

### Statistical analysis

Predicted probabilities (probability scores) were summarised using boxplots with overlaid individual observations.

Diagnostic performance was assessed using receiver operating characteristic (ROC) analysis. Ninety-five percent confidence intervals (95% CIs) for the area under the receiver operating characteristic curve (ROC-AUC) were determined using a bootstrap resampling method. The model predictions were resampled across 2,000 bootstrap iterations. The ROC-AUC was calculated for each iteration, and the lower and upper bounds of the 95% CI were defined as the 2.5 and 97.5 percentiles of the resulting bootstrap distribution, respectively. Sensitivity and specificity values were calculated using ROC-AUC analysis. Three sets of values were selected to indicate the utility of the diagnostic test in different clinical applications, for example, the sensitivity tuned model maintains a 90% cancer detection rate but results in higher false positive cases than a model tuned to maintain 90% specificity. Youden’s J Index was used to select the point at which the distance between the curve and the chance line (AUC = 0.5) is maximised. This point generally represents the optimal trade-off between sensitivity and specificity of a model.

Pre-processing and modelling were performed using Python 3.12, further analyses were performed using R version 4.6.0 and Python 3.12.

## Results

### Study cohort

A total of 146 participants were analysed.

### Distribution of predicted probabilities

The output of the machine learning model is a probability score which corresponds to the probability that an observation belongs to a specific class. **Figure 2** demonstrates clear separation between patients with bladder cancer and symptomatic controls using predicted probabilities. Similar distributions were observed for the spectra-only and spectra-plus-age models.

**Figure 2.**
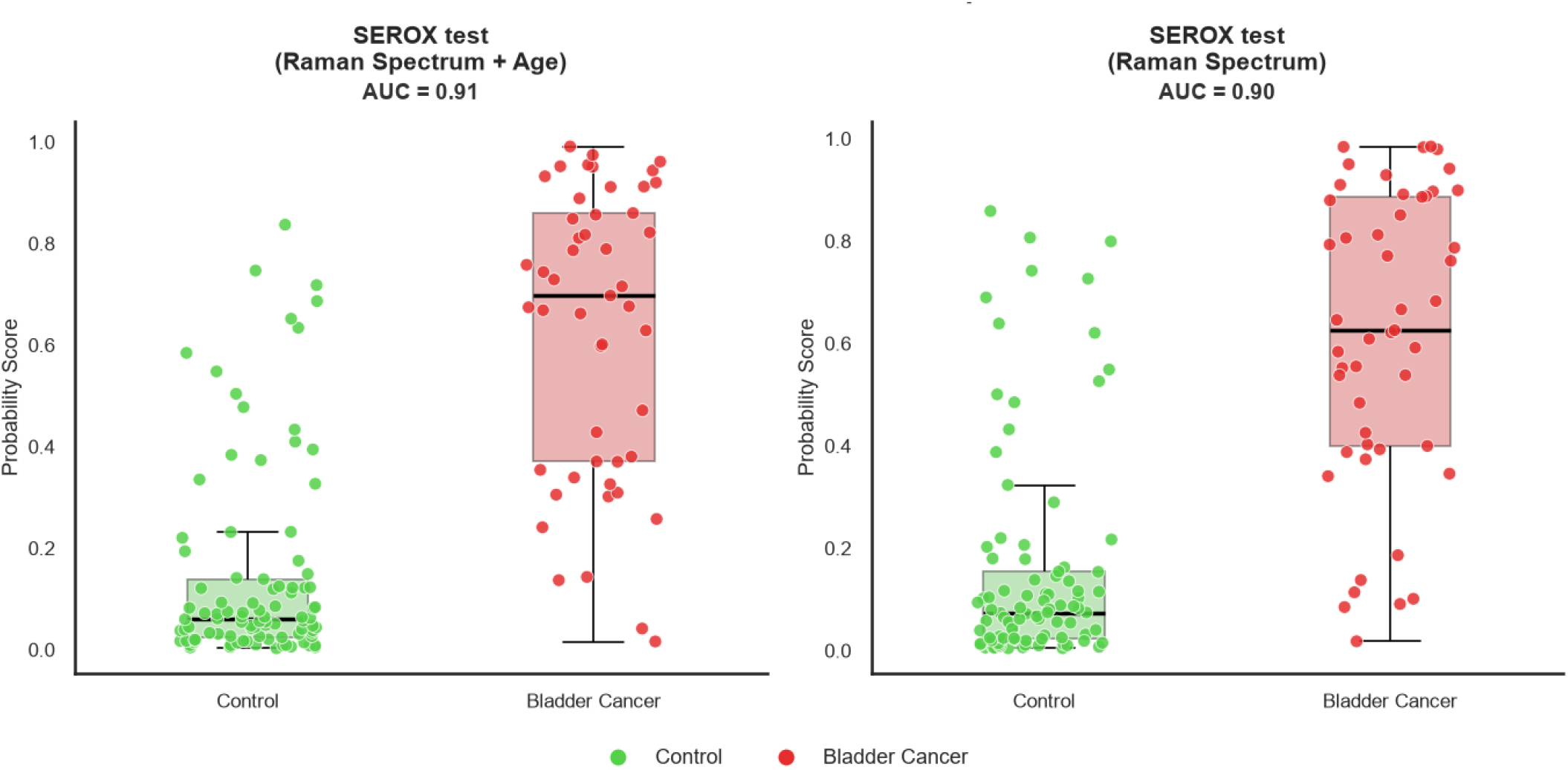
Boxplot of predicted probabilities of bladder cancer (red) vs. control (green) samples using **a)** Raman spectrum + Age model and **b)** Raman spectrum only model. Individual points represent the mean probability score for each sample obtained from 100 iterations of a 5-fold CV model. Boxplots represent the median and 25^th^ and 75^th^ percentiles of probabilities. Whiskers extend to the furthest data points within 1.5x IQR from the box edges, with outliers beyond this range shown as individual points.

### Diagnostic performance

Receiver operating characteristic curves were used to evaluate diagnostic performance of the machine learning models and are shown in **Figure 3**.

**Figure 3.**
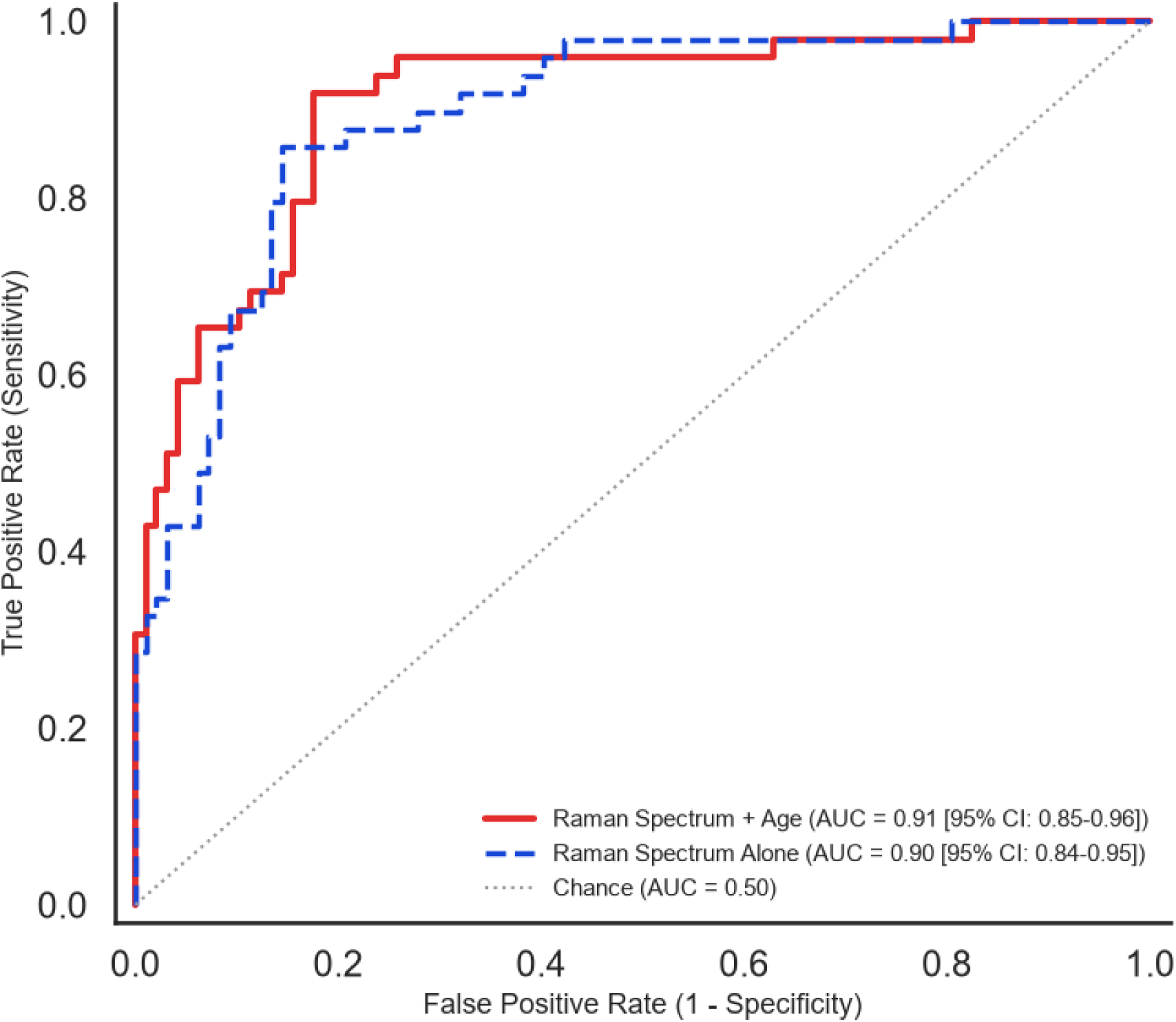
Receiver Operating Characteristic curve of Raman spectra + age model (red solid line, AUC 0.91 [95% CI 0.85-0.96]) and Raman spectra only model (blue dashed line, AUC 0.9 [95 % CI 0.84-0.95]).

The classifier based on Raman spectra alone achieved an AUC of approximately 0.90. Addition of age resulted in minimal further improvement (AUC approximately 0.91). Both models produced excellent diagnostic discrimination between bladder cancer samples and symptomatic controls. Sensitivity and specificity values for each model, at three different thresholds, are shown in Table 3.

**Table 3.**
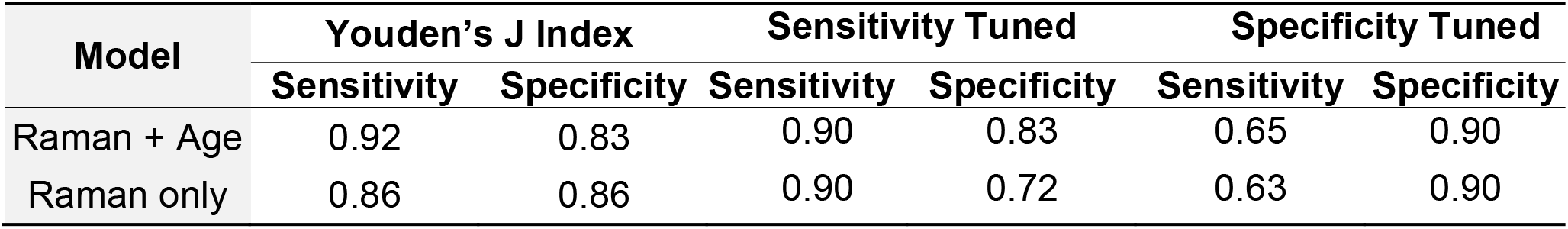
Sensitivity and specificity values, calculated from ROC analysis, at 3 different thresholds which suit different clinical applications.

## Discussion

The present study demonstrates that machine learning-assisted analysis of dried-serum Raman spectra can discriminate patients with bladder cancer from symptomatic controls with a high degree of diagnostic accuracy. The addition of age produced only minimal improvement, suggesting that the Raman-derived molecular signature contains most of the discriminatory information.

Current diagnostic pathways for bladder cancer rely heavily on cystoscopy. Although cystoscopy remains the diagnostic reference standard, it is invasive, resource-intensive and frequently performed in patients who ultimately prove not to have malignancy. [25]

[1] Consequently, there remains considerable interest in minimally invasive tests capable of improving patient selection before cystoscopy. [10] [7] [26] [8] Unlike targeted biomarker assays, Raman spectroscopy interrogates the global biochemical composition of serum rather than measuring predefined molecular targets. This approach may provide complementary biological information and offers the practical advantage that blood sampling is already embedded within many diagnostic pathways. [27] [28]

The diagnostic accuracy of the combined Raman spectroscopy and age model demonstrates strong clinical utility for a first-line bladder cancer triage within a symptomatic urology referral pathway. Using Youden’s J Index for ROC threshold optimisation, the model achieved 92% sensitivity and 83% specificity (Table 3), highlighting the clinical utility of this test as an initial non-invasive triage test. When used specifically as a rule-out triage tool tuned to maintain 90% sensitivity, a prerequisite for ensuring patient safety by minimising missed cancer cases, the test preserved an 83% specificity. In clinical practice, maintaining this level of specificity at high sensitivity provides a substantial reduction in unnecessary urgent cystoscopies for non-cancer patients, relieving pain-points in pathway referral whilst ensuring high-risk individuals are rapidly triaged to appropriate secondary care. Conversely, tuning the assay for high specificity (90%) results in a significant drop in sensitivity (65%), highlighting that the primary clinical strength of this serum-based approach lies as a high-sensitivity rule-out triage test.

The principal strengths of this study include the use of symptomatic controls and evaluation of a simple serum-based assay requiring minimal sample preparation. Additionally, 78% of bladder cancer cases correctly identified in this study were stage T1 or lower, indicating the potential use of the Raman based diagnostic for bladder cancer early detection. Three of the bladder cancer cases which were correctly identified as bladder cancer in this study were carcinoma *in situ* (CIS). CIS is often difficult to identify using white light guided cystoscopy and therefore the proposed Raman based diagnostic could support the earlier detection of more difficult to diagnose disease. The excellent discrimination observed in this retrospective cohort supports further prospective evaluation, in a larger clinical cohort.

Several limitations should also be acknowledged. This was a retrospective proof-of-concept study based on samples from a single biobank. External validation in independent cohorts will be required before clinical implementation can be considered. Although the present study evaluated bladder cancer, patients present with haematuria rather than with a diagnosis of bladder cancer. Future studies should therefore examine whether serum Raman spectroscopy can contribute to risk stratification across the wider spectrum of malignant causes of haematuria. Another limitation of this study is the low number of female samples available for analysis, particularly in the control cohort. Although urology clinic referrals tend towards 3:1 male to female and bladder cancer is more commonly diagnosed in males, future studies should aim to improve this class imbalance.

In conclusion, serum Raman spectroscopy represents a promising approach to minimally invasive bladder cancer detection. Prospective studies should determine whether this technology can improve patient triage while reducing unnecessary invasive investigations.

## Data Availability

The processed model outputs underlying Figures 2 and 3 (per-sample mean predicted probabilities with class labels and age) may be made available from the corresponding author on reasonable request, subject to approval by the QUANTUM Research Tissue Bank and a data-sharing agreement.

## List of abbreviations

AUC: Area under curve
CIS: carcinoma in situ
CV: cross validated
IQR: interquartile range
PCA: principal component analysis
PUNLMP: Papillary urothelial neoplasm of low malignant potential
ROC: Receiver operator characteristic curve
XGBoost: eXtreme Gradient Boosting

## Declarations

## Acknowledgments

This research has been conducted with support from the QUANTUM (QUAlity biobanking iNvesTigating Urological Malignancies) Research Tissue Bank, which is operated by the University of Oxford.

We acknowledge the contribution to this study made by the Oxford Centre for Histopathology Research and the Oxford Radcliffe Biobank, which are funded by the University of Oxford, the Oxford CRUK Cancer Centre, and the NIHR South Central RRDN.

## Author contributions

Conceptualisation: CM, KM, IS, AF, NFA. Methodology: KM, NFA, AF. Machine learning analysis: AF, BS. Writing: IS, KM, BS. Writing, reviewing and editing: All authors.

### Funding

No public funding was used in this study.

### Ethics approval and consent to participate

Ethical approval for this study was obtained from the relevant UK NHS Research Ethics Committees under REC references 24/SC/0220 (Oxford Radcliffe Biobank) and 22/NS/0004 (QUANTUM Biobank).

This study was performed under the ethics approval granted by South Central - Oxford C Research Ethics Committee to Oxford Radcliffe Biobank on 15 October 2024 (OCHRe ref: 26/A007, REC reference 24/SC/0220, IRAS project ID: 340834). The Committee confirmed that the favourable ethical opinion applies to all research projects conducted in the UK using tissue or data supplied by the tissue bank, provided that the release of tissue or data complies with the stated conditions; and that it is not be necessary for these researchers to make project-based applications for ethical approval, as they will be deemed to have ethical approval from this committee.

Experiments were performed in accordance with the World Medical Association Declaration of Helsinki in a facility equipped to Biosafety Level 2. The laboratory has an HTA licence (12811) and is ISO 20387:2018 (Biobanking) accredited (31342).

### Consent for publication

Not applicable

